# Global metabolite profiling in feces, serum, and urine yields insights into energy balance phenotypes induced by diet-driven microbiome remodeling

**DOI:** 10.1101/2025.02.05.25321733

**Authors:** Daria Igudesman, GongXin Yu, Tumpa Dutta, Elvis A. Carnero, Rosa Krajmalnik-Brown, Steven R. Smith, Karen D. Corbin

## Abstract

**Background.:** Preclinical literature and behavioral human data suggest that diet profoundly impacts the human gut microbiome and energy absorption—a key determinant of energy balance. To determine whether these associations are causal, domiciled controlled feeding studies with precise measurements of dietary intake and energy balance are needed. Metabolomics—a functional readout of microbiome modulation—can help identify putative mechanisms mediating these effects. We previously demonstrated that a high-fiber, minimally processed Microbiome Enhancer Diet (MBD) fed at energy balance decreased energy absorption and increased microbial biomass relative to a calorie-matched fiber-poor, highly processed Western Diet (WD).

**Objective.:** To identify metabolic signatures distinguishing MBD from WD feeding and potential metabolomic mechanisms mediating the MBD-induced negative energy balance.

**Methods.:** We deployed global metabolomics in feces, serum, and urine using samples collected at the end of a randomized crossover controlled feeding trial delivering 22 days of an MBD and a WD to 17 persons without obesity. Samples were collected while participants were domiciled on a metabolic ward and analyzed using Ultrahigh Performance Liquid Chromatography-Tandem Mass Spectroscopy. Linear mixed effects models tested metabolite changes by diet. Weighted gene network correlation analysis identified metabolite modules correlated with energy balance phenotypes.

**Results.:** Numerous metabolites consistently altered in the feces, fasting serum, and/or urine may serve as putative dietary biomarkers of MBD feeding. Fecal diet-microbiota co-metabolites decreased by an MBD correlated with reduced energy absorption and increased microbial biomass. An MBD shifted the urinary metabolome from sugar degradation to ketogenesis—evidence of negative energy balance.

**Conclusions.:** Precisely controlled diets disparate in microbiota-accessible substrates led to distinct metabolomic signatures in feces, fasting serum, and/or urine. These diet-microbiota co-metabolites may be biomarkers of a “fed” (MBD) or “starved” (WD) gut microbiota associated with energy balance. These findings lay the foundation for unveiling causal pathways linking diet-microbiota co-metabolism to energy absorption.

## Introduction

The complex interplay between diet, the gut microbiota, and host physiology is a primary target for addressing the obesity epidemic and its core underpinning of sustained positive energy balance. Intriguing observational and preclinical studies have linked diet-gut microbiota interactions to energy balance phenotypes (2), but progress towards developing diet-focused microbiota therapeutics for obesity has been stilted by a dearth of causal evidence in humans. Human controlled feeding studies are a key tool that can facilitate the discovery of mechanisms and biomarkers by which host and gut microbiota coordinately alter energy balance (3, 4). Accordingly, we employed a tightly controlled feeding paradigm to deliver two eucaloric and isocaloric diets: 1) a Microbiome Enhancer diet (MBD) high in fiber and resistant starch, with large particles of food (whole foods) to increase the delivery of substrate to the colon, and limited in highly processed foods, and 2) a macronutrient-matched Western diet (WD) devoid of microbiota-accessible carbohydrates (5) (NCT02939703). Critically, we used precise and comprehensive methods to measure energy balance. We found that an MBD induced a 116 kcal/day *excess* fecal energy loss relative to a WD with no difference in energy expenditure (5). This excess fecal energy loss corresponded to a mean ∼5% decrease in metabolizable energy (referred to as “energy absorption” throughout this manuscript), which is approximated by the proportion of dietary energy available to the human host after accounting for fecal energy loss (6). The decrease in energy absorption was not just due to the expected lower digestibility of foods, but also due to the shunting of energy towards growth of bacterial biomass (5). Although it was not significantly altered between diet conditions, colonic transit time (CTT) was on average ∼10 hours faster on an MBD, providing less time for colonic energy extraction (5). An MBD also increased circulating levels of the satiety hormone pancreatic polypeptide (PP), foreshadowing the potential of an MBD to stave off hunger during longer-term negative energy balance. These four negative energy phenotypes could be part of a comprehensive strategy of small changes for achieving and maintaining weight loss (7).

Metabolites are key mediators on the causal pathway between diet, the gut microbiota, and energy balance (8). Specifically, gut microbial metabolites reflect the harvesting of dietary energy from nondigestible food substrates, promote the secretion of satiety hormones, and perform signaling functions regulating fat oxidation, as in the case of short-chain fatty acids (9, 10). Most mechanistic evidence linking diet-gut microbiota co-metabolism to energy balance is restricted to animal models (11). To unravel causal pathways in humans amenable to targeted dietary interventions, robust biomarkers of these processes are needed. Thus, the objectives of this exploratory analysis were twofold. First, we sought to identify potential diet-microbiota biomarkers of MBD vs. WD feeding using global metabolomics in the feces, fasting serum, and urine. We did so using a combination of principal coordinate analysis and differential abundance testing. Secondly, we aimed to generate novel mechanistic hypotheses from the host and microbial phenotypes of energy balance we observed on an MBD. In this step, rather than cataloguing all metabolite changes that were induced by our controlled diets, we explored links between differentially abundant metabolites and the energy balance related phenotypes of reduced energy absorption, increased microbial biomass, reduced CTT, and increased circulating PP we observed on an MBD relative to a WD using network analysis. We believe our approach to be a useful starting point for pinpointing metabolic pathways through which diet-microbiome interactions promote negative energy balance and could be exploited as part of a comprehensive strategy to curb the obesity epidemic.

## Methods

### Study participants

Participant recruitment and characteristics have been described in detail in both our published study protocol (12) and the topline results manuscript (5), with a participant flow diagram included in the latter. Briefly, we recruited generally healthy men and women aged 18-45 with a body mass index (BMI) ≤30 kg/m^2^ (all participants had a BMI within the normal to overweight range) between the years 2017 and 2019. OIder adults, individuals with obesity or chronic health conditions, and those who had recently taken antibiotics were excluded to minimize confounding effects of these characteristics on the gut microbiota. A full list of inclusion and exclusion criteria are published in the topline manuscript (5). Participants provided informed consent, and the research was conducted in compliance with all applicable ethical and institutional research requirements. The study was approved by the AdventHealth Institutional Review board (Orlando, FL, USA).

### Study design

The design of our randomized crossover controlled feeding study has been described extensively (5, 12). The general design is shown in **Fig. 1**. Briefly, participants underwent two dietary intervention periods that were 22 days each, separated by a >14-day washout period. The first 11 days of each period were outpatient with uneaten foods weighed back by our metabolic kitchen, and the second 11 days were domiciled in our clinic research unit and whole-room indirect calorimeters. Participants were fed to maintain their baseline body weight.

**Fig. 1:**
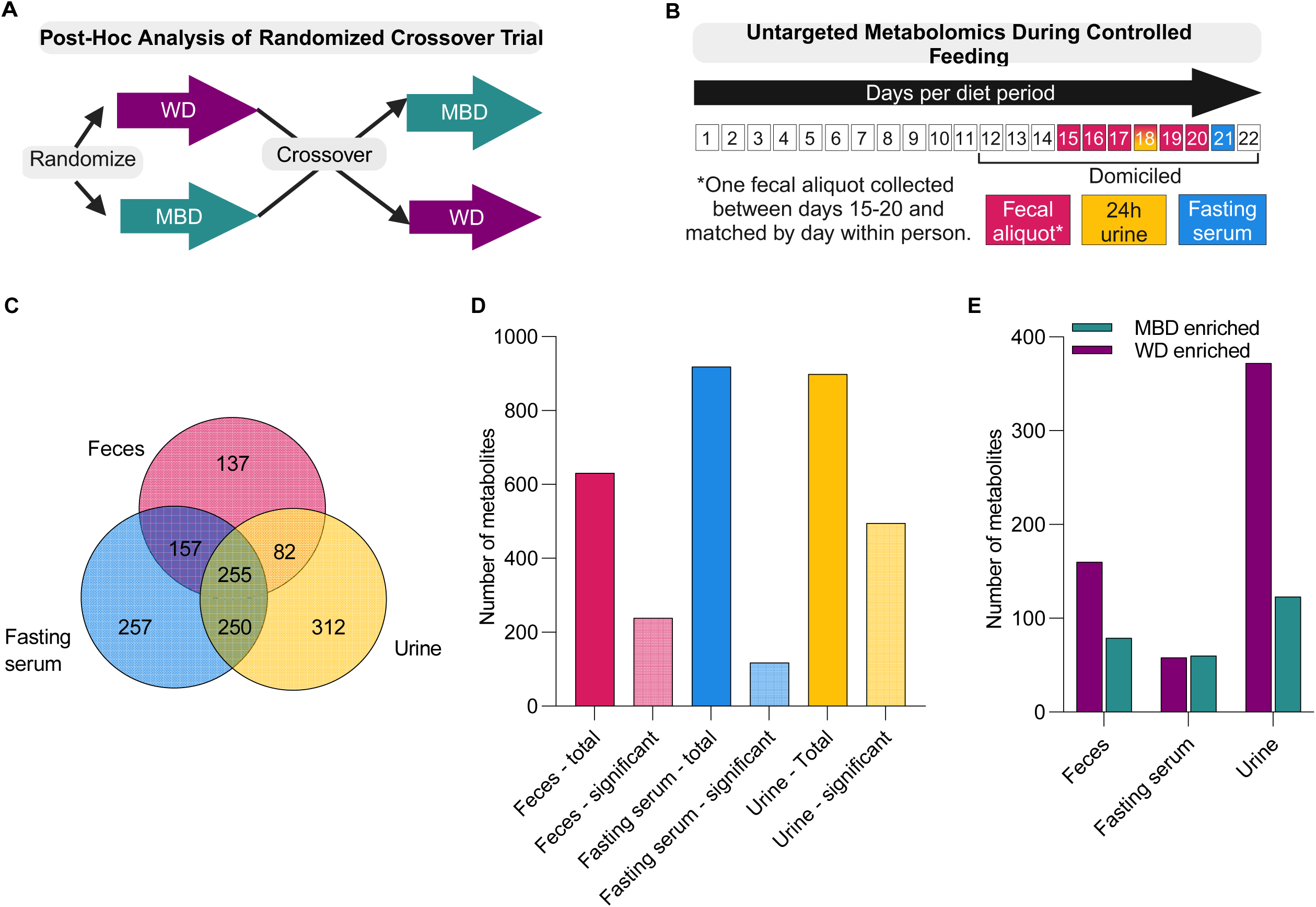
A Microbiome Enhancer Diet generates unique metabolite signatures of dietary microbiome modulation. **A.** Schematic of randomized crossover design. **B.** Sequence of metabolomics measurements during each 22-day controlled feeding period. **C.** Retained metabolites in feces, fasting serum, and urine after data filtration. **D.** Total and significant metabolites. **E.** Number of significant metabolites enriched by an MBD or WD. Results in **C-D** based upon limma linear mixed models with participant as random effect and diet, period, and sequence as fixed effects. Results in **C-E** generated from N=17 participants for feces and urine, and N=15 participants for fasting serum. MBD—Microbiome Enhancer Diet; WD—Western Diet.

### Dietary intervention

The MBD was designed to maximize the delivery of nondigestible food components to the colon where it would be available for microbial degradation, whereas a WD aimed to maximize the digestibility of foods in the upper GI tract. The four dietary drivers of microbial remodeling were dietary fiber, large food particle size, resistant starch, and minimal processing, as previously described (12). Diets were matched for calories, macronutrients, and types of foods wherever possible. Details about how diets were prepared, provided, and chemically analyzed are outlined in our previous publications (5, 12).

### Sample collection, processing and shipment

Blood, urine, and fecal samples were collected in our metabolic research unit using standardized protocols (AdventHealth Translational Research Institute, Orlando, FL, USA). Fecal samples were processed within an hour of collection under an anaerobic hood and held on ice during processing, after which the aliquots used for metabolomics analyses were snap frozen without additives and stored at −80° C. The fecal aliquot used for measurement of global metabolomics was taken between days 15-19 of controlled feeding during each diet period, with the day of collection matched as closely as possible within a participant. Fasting serum for metabolomics analysis was collected at approximately 8:45 am (15 minutes prior to breakfast) on day 20 of controlled feeding in each diet condition. 24-hour urine samples were collected on day 18 of each controlled feeding period. Serum, urine, and fecal samples were sent to Metabolon for measurement of untargeted metabolomics. Upon receipt, samples were stored at -80°C until analyzed.

### Metabolizable energy

Calculation of metabolizable energy (i.e., energy absorption) is described in depth in our design and topline manuscripts (5, 12). Briefly, chemical oxygen demand, a unit of energy employed by microbial ecologists, was measured via chemical analysis in feces and converted to fecal kilocalories using our published mathematical model (13). We normalized chemical oxygen demand in composite fecal samples collected over six days of domiciled controlled feeding to recovery of polyethylene glycol, a fecal marker that allows for quantification of fecal substrates once participants reach a steady state, as previously described (5).

### Microbial biomass

We used quantitative PCR of the bacterial 16S rRNA gene to estimate gene copy number, as previously described (5).

### Colonic transit time

A radiotransmitter motility capsule was used to determine colonic transit time between days 15-19 of each diet condition, with the day of capsule ingestion matched as closely as possible within a participant (SmartPill™; Medtronic, Minneapolis, MN). The SmartPill™ was administered immediately after breakfast, as previously described, with colonic transit times reported elsewhere (5).

### Pancreatic polypeptide

Plasma PP was evaluated at 18 nominal timepoints before and after the breakfast, lunch, and dinner meals over the course of 12 hours (between 8:30am and 8:30pm) with V-PLEX Metabolic Panel 1 Human Kit (MesoScale Diagnostics, Rockville, MD; Product # K15325D). Timepoints were taken relative to each meal, at −30, −15, +30, +60, + 120, and +180min. The two fasting measurements were averaged and used in the network analysis correlating fasting metabolite levels with fasting PP. For network analysis using fecal and urine data, all 18 PP measurements were used to calculate an area under the curve using the trapezoidal rule (14). The meals provided during these 12 hours included a standardized breakfast consisting of a standardized liquid meal and a lunch and dinner meal consistent with the assigned diet.

### Untargeted metabolomics

Sample preparation and quality control methods have previously been described in detail, as have the mass spectroscopy analytic methods (15) (Metabolon, Inc., Durham, NC). Briefly, fecal samples were lyophilized prior to metabolomics analysis. All methods utilized a Waters ACQUITY Ultrahigh Performance Liquid Chromatograph (UPLC) connected to a Thermo Scientific Q-Exactive high resolution/accurate mass spectrometer interfaced with a heated electrospray ionization source. The Orbitrap mass analyzer was operated at 35,000 mass resolution. The MS analysis alternated between MS and data-dependent MSn scans using dynamic exclusion. The scan range varied slightly between methods but covered 70-1000 m/z. Raw data was extracted, peak-identified and processed using Metabolon’s proprietary software. Compounds were identified by comparison to library entries of purified standards or recurrent unknown entities. The metabolite library is based on authenticated standards that contains the retention time/index, mass to charge ratio (*m/z)*, and chromatographic data (including MS/MS spectral data) on all molecules present in the library. More than 3300 commercially available purified standard compounds have been acquired and registered into Laboratory Information Management System for analysis on all platforms for determination of their analytical characteristics. Only annotated metabolites were reported.

#### Data curation

The quality control and curation processes were designed to ensure accurate and consistent identification of true chemical entities, and to remove those representing system artifacts, mis-assignments, and background noise. Proprietary visualization and interpretation software are used to confirm the consistency of peak identification among the various samples. Library matches for each compound were checked for each sample and corrected if necessary.

#### Metabolite Quantification and Data Normalization

Metabolite peaks were identified, extracted, and quantified then subjected to quality control steps prior to normalization in terms of raw area counts (15). Peaks were quantified using area-under-the-curve. Between 733 and 2398 mg feces, 100 µL serum, and 1000 µL urine were used for metabolomics analysis. For fecal and urine samples, biochemical data were normalized to fecal mass and 24-hour urine volume, respectively, to account for differences in metabolite levels due to differences in the amount of material present in each sample.

#### Filtering and imputing missing data

Appropriate to our randomized crossover design, metabolites present in ≥75% of samples on at least one diet were retained to avoid losing important information about metabolites depleted on one diet but not the other. Missing values were then imputed with the minimum value per metabolite, as previously published (16).

### Bioinformatics and data visualization

The effects of diet on global metabolite profiles were first visualized using principal components analysis (PCA) and volcano plots in Graphpad Prism 9. Principal components were computed using median-scaled, imputed, and normalized (in the case of feces and urine) values that were log_2_ transformed and scale-standardized prior to PCA. Parallel PCA analysis was employed to visualize separation of metabolite values by diet according to the first two principal components; this PCA method is recommended by Graphpad and selects principal components based on a comparison of input data to 1,000 iterations of simulated noise (17). The R package limma (18, 19) version 3.62.1 was used to test for differentially abundant metabolites using a linear mixed effects model specifying participant as a random effect and treatment, period, and sequence as fixed effects. This analytic method makes no assumptions about prior probabilities (i.e., the proportion of differentially abundant metabolites) (18, 19). The Benjamin-Hochberg method was used to correct the limma p-values for the false discovery rate. Heatmaps of significantly altered metabolites were created using the pheatmap package in R studio version 2024.04.2, with log_2_ transformed metabolites. Variable importance plots were created using the randomForest R package version 4.7-1.2 and visualized using the package ggplot2. Figures showing commonly altered metabolites in the feces and fasting serum were created using the R package ggplot2 version 3.5.1.

Network analysis was performed specifically on fecal metabolomics data using the R package WGCNA (20) version 1.73, which first constructs a weighted co-expression network analysis with clusters of highly correlated metabolites (i.e., modules). Given the crossover design, modules were constructed based on the log_2_-transformed metabolite ratios (MBD/WD) to avoid violating the assumption of independent observations (i.e., including the same participant’s data twice in correlation analyses). As this analysis is exploratory and hypothesis generating, we included only the fecal metabolites that were differentially abundant between an MBD and a WD based on a p-value threshold of <0.05 according to the limma analysis to reduce the possibility of spurious findings. Module eigengenes (i.e., the first principal component of a module representing the constituent metabolite abundance profiles (20)) and individual metabolites within each module were then correlated with log_2_-transformed ratios (MBD/WD) in energy balance phenotypes (i.e., metabolizable energy, microbial biomass, colonic transit time, and pancreatic polypeptide) using Pearson correlation. Correlations between network modules and energy balance phenotypes were visualized using the labeledHeatmap function within the WGCNA package. The cuttree function within WGCNA was used to identify the presence of outlier samples, with no outliers detected.

### Statistics

Descriptive statistics for continuous variables are presented as mean ± standard deviation. Pearson correlations between log_2_ fold changes in metabolites and log_2_ fold changes in energy balance phenotypes were visualized in Graphpad Prism 9.

## Results

### A Microbiome Enhancer Diet generates unique metabolite signatures of dietary microbiome remodeling

We fed two controlled diets in random order to 17 generally healthy participants aged 30.8 ± 1.9 years (body mass index 25.1 ± 0.52 kg/m^2;^ **Supplemental Figure 1**). The study sample was representative of the Orlando Metro area, with n=11 (64.7%) reporting a Black race (n=1 Unknown, n=5 White race), and n=6 (35.3%) reporting a Hispanic or Latino ethnicity. Participant characteristics have previously been described in detail (5). The two diets were a fiber-rich, whole food, high resistant starch MBD (26.0 ± 0.06 g fiber/1,000 calories) and a fiber-depleted, highly processed WD (6.4 ± 0.02 g fiber/1,000 calories) (5). The negative energy balance (i.e., excess fecal energy loss) induced by an MBD relative to a calorie-matched WD suggested a gap in existing paradigms for determining dietary energy availability (12), which do not account for the role of the gut microbiota. To explore potential mechanisms underlying the negative energy balance induced by dietary microbiome remodeling, we performed untargeted metabolomics of the feces, fasting serum, and 24-hour urine (5). Samples were collected following 15-20 days of controlled feeding of an MBD and a WD in random order, with each of 17 participants serving as their own control (**Figure 1A**). The first 11 days of controlled feeding were outpatient. Participants were domiciled in the metabolic ward in the final 11 days of controlled feeding when all study measurements including metabolomics samples were collected (**Figure 1B**). Participants consumed virtually 100 percent of calories provided (5). Metabolites were measured using Ultrahigh Performance Liquid Chromatography-Tandem Mass Spectroscopy (UPLC-MS/MS) following standard procedures (15). Specific to our dataset, we retained metabolites present in ≥75% of samples on at least one diet as the data filtering criterion, as several metabolites were largely depleted by one diet but not the other. The phenomenon of the near absence of certain metabolites on one diet but not the other was expected based on the stark differences in microbiota accessible carbohydrates between diets. After filtering the data, 631 of 822 fecal metabolites, 919 of 1049 fasting serum metabolites, and 899 of 963 urinary metabolites progressed to the statistical analysis (number of retained metabolites shown in **Figure 1C**). Using analytic procedures appropriate to our randomized crossover design with adjustment for the false discovery rate, 239 fecal metabolites, 118 fasting serum metabolites, and 495 urinary metabolites were altered between diet conditions (q<0.05, **Figure 1D**; **Supplemental Table 1**). In the feces, 79 metabolites were increased and 160 were decreased by MBD compared to WD feeding (**Figure 1E**). In the fasting serum, 60 metabolites were increased and 58 metabolites were decreased by MBD relative to WD feeding. In the urine, 372 metabolites were decreased and 123 metabolites were increased by MBD relative to WD feeding. Next, we pinpointed how alterations in fecal metabolite superpathways (i.e., amino acids, carbohydrates, cofactors and vitamins, energy, lipids, peptides, nucleotides, and xenobiotics) and the constituent metabolites within each superpathway might relate to microbiome-mediated changes in host energy absorption.

### A Microbiome Enhancer Diet decreases fecal metabolic byproducts of energy and carbohydrates in tandem with reduced host energy absorption

Here, we sought to explore potential mechanisms underlying the reduced host energy absorption induced by the interaction of host, diet, and microbes on an MBD. We did so by characterizing the corresponding shift in global fecal metabolomic signatures induced by our two diets high and low in microbiota-accessible diet-derived substrates. A clear separation of fecal metabolite profiles by diet was apparent according to the principal components analysis (**Figure 2A**; **Supplemental Figure 2A**). Our analysis revealed that the 239 differentially abundant fecal metabolites (**Figure 2B**) spanned eight metabolite superpathways (**Figure 2C**). Indeed, fecal amino acid fermentation byproducts including the microbiota metabolite P-cresol (log_2_FC -1.6, q=5.81E^-05^; **Figure 2D**), isovalerate (C5) (log_2_FC -1.2, q=0.0001; **Figure 2E**), valerate (5:0) (log_2_FC -0.80, q=0.02), and phenylacetate (log_2_FC -0.71, q=0.03) were decreased by an MBD. Increased byproducts of amino acid fermentation (i.e., p-cresol) under conditions of fiber scarcity are posited to contribute to the positive energy balance phenotype of obesity (21). On the other hand, there was a unanimous increase in the microbial polyphenol fermentation byproduct Urolithin A (5) by an MBD. This metabolite showed the largest increase during MBD relative to WD feeding (log_2_FC 3.9, q=0.004; **Figure 2F**) and has been associated negatively with central obesity (22). Whether the underlying mechanism is related to host energy absorption remains to be determined.

**Fig. 2:**
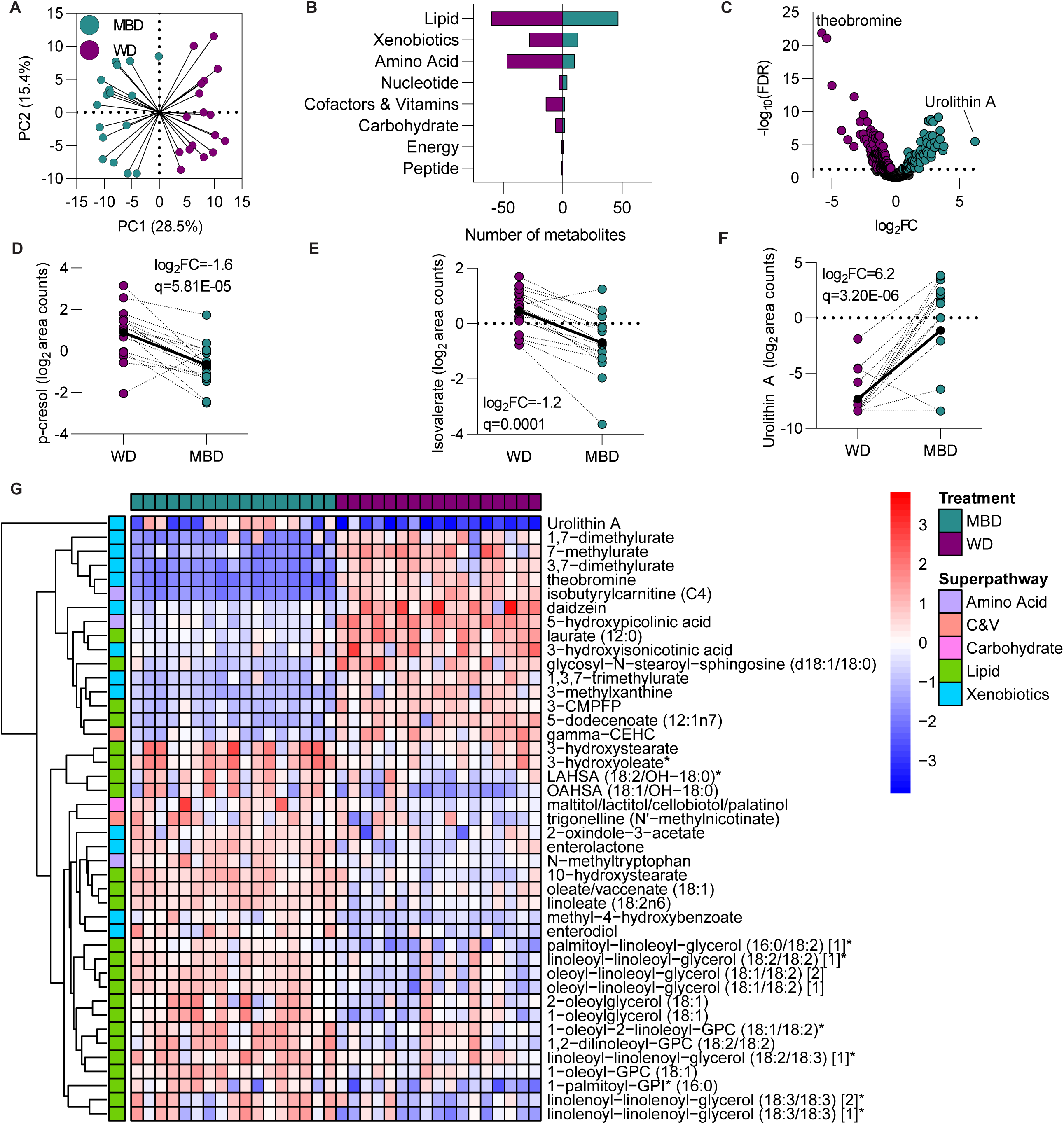
A Microbiome Enhancer Diet decreases fecal metabolic byproducts of energy and carbohydrates in tandem with negative host energy balance phenotypes. **A.** Principal components analysis of the 239 fecal metabolites that significantly varied by diet. **B.** Metabolite differences within superpathways. Negative counts indicate the number of metabolites decreased by an MBD relative to a WD for each superpathway. **C.** Volcano plot of metabolite log_2_ foldchanges against -log_10_ q-value. Log_2_ area counts of fecal p-cresol (**D.**), isovalerate (**E.**), and Urolithin A (**F.**) by diet; log_2_ foldchanges and p-values are from limma mixed effects models. **G.** Fecal metabolites that were significantly different by diet with an absolute log_2_FC ≥2 (q<0.05). Metabolites included in **A-G** based upon limma linear mixed models with participant as random effect and diet, period, and sequence as fixed effects. N=17 participants for all panels. Metabolite names ending in ‘*’ or ‘**’ indicate compounds for which a standard is not available but for which there is confidence in the compound identity (70). 3-CMPFP—3-carboxy-4-methyl-5-pentyl-2-furanpropionate; C&V—Cofactors and vitamins; PC—principal coordinate; MBD—Microbiome Enhancer Diet; WD—Western Diet.

In addition to amino acids, fecal metabolites within the energy and carbohydrate superpathways were decreased by MBD feeding, potentially owing to decreased simple sugar intake and increased undigestible fiber intake on an MBD. Such dietary conditions create an environment for enhanced microbial fermentative capacity and biomass growth—correlates of reduced host energy absorption (5). In line with this principle were MBD-induced decreases in the carbohydrate metabolites N-acetylglucosamine/N-acetylgalactosamine (log_2_FC -1.36, q=0.0005) and N-acetylneuraminate (log_2_FC -0.95, q=0.008), which promote the growth of mucin-degrading bacteria (23–26). Decreases in these two metabolites could serve as biomarkers of a microbial biomass that is allowed to expand when adequately provided with fibrous substrates, thus promoting reduced host energy absorption. Similarly, an MBD decreased fecal levels of citraconate/glutaconate within the energy metabolism superpathway/TCA cycle subpathway (log_2_FC -0.86, q=0.0002) and the monosaccharide fucose (log_2_FC -1.08, q=6.02E^-05^). Although it is a SCFA precursor, fucose decreases microbial biomass because it is an inefficient energy source for gut microbes (27), and could therefore be an indicator of increased host energy absorption. Despite global decreases in carbohydrate metabolites, an MBD increased fecal alpha-ketoglutarate, a central regulator of the TCA cycle and an energy source for the gastrointestinal tract (28) (log_2_FC 0.59, q=0.04). Collectively, we hypothesize that an MBD produces a negative host energy balance (5) through fecal metabolite changes that augment microbial biomass growth and thereby decrease host energy absorption.

We found striking alterations in fecal lipid metabolites on an MBD (**Figure 2G**). Lipid classes implicated in obesity and positive energy balance, including ceramides, sphingosines, secondary bile acids, and the saturated fatty acids palmitate (16:0) and myristate (14:0), were decreased during MBD relative to WD feeding. Although lipids are not readily available as an energy source for the gut microbiota, these lipids may be structural components of microbial biomass (29). Given the vast enzymatic capabilities of gut microbes to chemically modify lipid moieties (29), this finding may also be indicative of an altered microbial ecosystem with a differential capacity to modify host and dietary lipids (5). These fecal lipid metabolites may therefore serve as potential biomarkers of MBD-induced reductions in host energy absorption promoted by an altered microbial ecology.

A complementary approach to identifying fecal metabolomic signatures of our controlled diets that differentially impacted energy balance is to ask which metabolites are key for the classification of diet assignment. To achieve this, we deployed a random forest machine learning model predicting diet condition (MBD or WD) from the 239 differentially abundant fecal metabolites. These 239 metabolites classified diet with 100% accuracy (**Supplemental Figure 2B**). Enterolactone, a microbial metabolite of plant lignan degradation (30), was the top MBD-enriched fecal metabolite contributing to diet classification accuracy. Enterolactone has been inversely associated with obesity in a population-based study (31). Thus, random Forest machine learning revealed additional potential metabolite mediators by which an MBD may reduce host energy absorption.

To consolidate metabolite alterations into potential metabolic mechanisms underlying the MBD-induced negative energy balance, we performed pathway analysis on the differentially abundant fecal metabolites (q<0.05). The top MBD-depleted pathway was branched-chain amino acid catabolism (z-score –1.4, p=1.17E^-05^; **Supplemental Figure 3**), as predicted by increased alpha-ketoglutarate and glutamate, and decreased 3-methyl-2-oxovalerate, 4-methyl-2-oxopentanoate, 3-methyl-2-oxobutyrate, isoleucine, leucine, and valine. Meanwhile, the top MBD-enriched pathway was PPARα (z-score 0.45, p=6.95E^-05^), as predicted by increased linoleate (18:2n6) and decreased bilirubin, deoxycholate, lithocholate, and palmitate (16:0). PPARα has been shown to increase satiety and reduce body weight in both rodent models (32) and in humans (33), which may be mediated by the gut microbiota (34). These results may reflect that insufficient dietary fiber necessitates amino acid catabolism by gut microbes. Thus, fecal metabolites indicative of decreased amino acid catabolism may be biomarkers of an increased microbial biomass leading to reduced host energy absorption on an MBD.

### Networks of fecal host-microbial co-metabolites correlate with diet-induced alterations in energy balance phenotypes

We next explored the hypothesis that fecal metabolomic signatures altered differentially between diets are correlated with negative energy balance phenotypes, including reduced host energy absorption. We did so by using weighted gene correlation network analysis (WGCNA) (20) to construct modules of intercorrelated metabolites in an unbiased manner. Given our randomized crossover repeated measures design, the modules were constructed using log_2_ fold-changes in metabolites comparing an MBD to a WD within each participant. We correlated the resultant network modules and individual metabolites within each module to the changes in negative energy balance phenotypes. These phenotypes included metabolizable energy (i.e., host energy absorption), microbial biomass (16S qPCR), CTT, and circulating PP measured over 12 hours (incremental area under the curve).

Global fecal metabolites that were found to be perturbed by diet before multiple hypothesis testing correction (282 metabolites, p<0.05) clustered within one of eight metabolite modules (**Figure 3A**). Module-trait correlations revealed that none of the fecal modules were correlated with CTT or PP. However, the turquoise module comprised of 80 metabolites was correlated with increased energy absorption (Pearson’s r=0.61, p=0.01), and tended to be correlated with decreased microbial biomass (r=-0.44, p=0.07). Thus, the turquoise module may represent an interacting combination of biomarkers or mediators of positive energy balance. The turquoise module metabolites that were commonly correlated with increased energy absorption and decreased biomass were the branched-chain amino acid derivatives N-acetylvaline (**Figure 3B-C**) and beta-hydroxyisovalerate (**Figure 3D-E**), the intermediary nicotinamide (NAD) precursor (35) quinolinate (**Figure 3F-G**), and the pro-inflammatory bacterial sphingolipid, phytosphingosine (35) (**Figure 3H-I**). Of note, these correlations were robust to removal of one statistical outlier for change in energy absorption (p<0.1), and all of the corresponding metabolites were decreased after MBD relative to WD feeding. These results lead us to hypothesize that amino acid byproducts of protein catabolism generated under conditions of low fiber intake may hamper microbial biomass growth or are biomarkers of this phenomenon (36), reflecting greater host energy absorption. Whether the metabolites we identified have mechanistic importance for energy balance or are simply derivatives of an altered microbial biomass requires confirmation in future research.

**Fig. 3:**
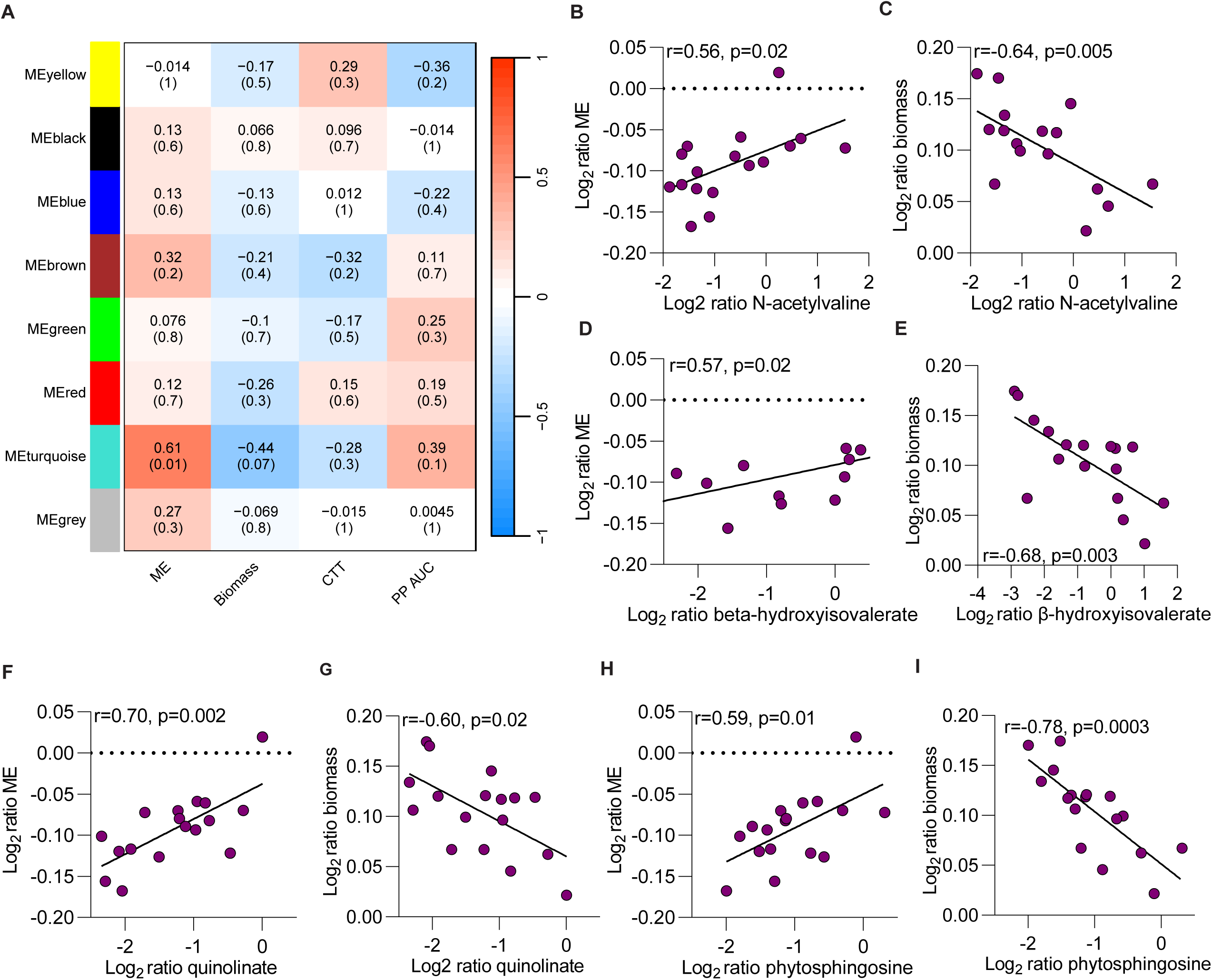
Networks of fecal host-microbial co-metabolites correlate with diet-induced alterations in energy balance components. **A.** Metabolite-module Pearson correlations (WGCNA). **B-I.** Pearson correlations of change in turquoise module metabolites correlated with change in metabolizable energy (i.e., energy absorption) and biomass; dots are purple as they were decreased by MBD relative to WD feeding (i.e., higher on WD). The 282 included metabolites (**A.**) were differentially abundant by diet (p<0.05) according to upon limma linear mixed models with participant as random effect and diet, period, and sequence as fixed effects. N=17 participants for all results, except N=15 for correlations with PP AUC in (**A.**). C&V— Cofactors and vitamins; PC—principal coordinate; MBD—Microbiome Enhancer Diet; WD—Western Diet; ME—module eigengene (**A.**) or metabolizable energy (**C-I.**).

### Host-diet-microbiome interactions characterize a circulating metabolome that distinguishes an MBD from a WD

Evaluating snapshots of metabolite profiles in circulation at specific timepoints can reveal the more complex landscape of metabolic interactions between host and microbes. Thus, we next sought to understand whether circulating small molecules altered by our controlled diets could reveal a biomarker signature of the host-diet-microbiome interactions influencing energy balance. A principal components analysis revealed that the 118 fasting serum metabolites that differed by diet (q<0.05) drove a clear separation of metabolomic signatures (**Figure 4A**). Unlike the fecal data, including all 919 retained fasting serum metabolites, including those that did not differ by diet, did *not* reveal a diet separation (**Supplemental Figure 4A**).

**Fig. 4:**
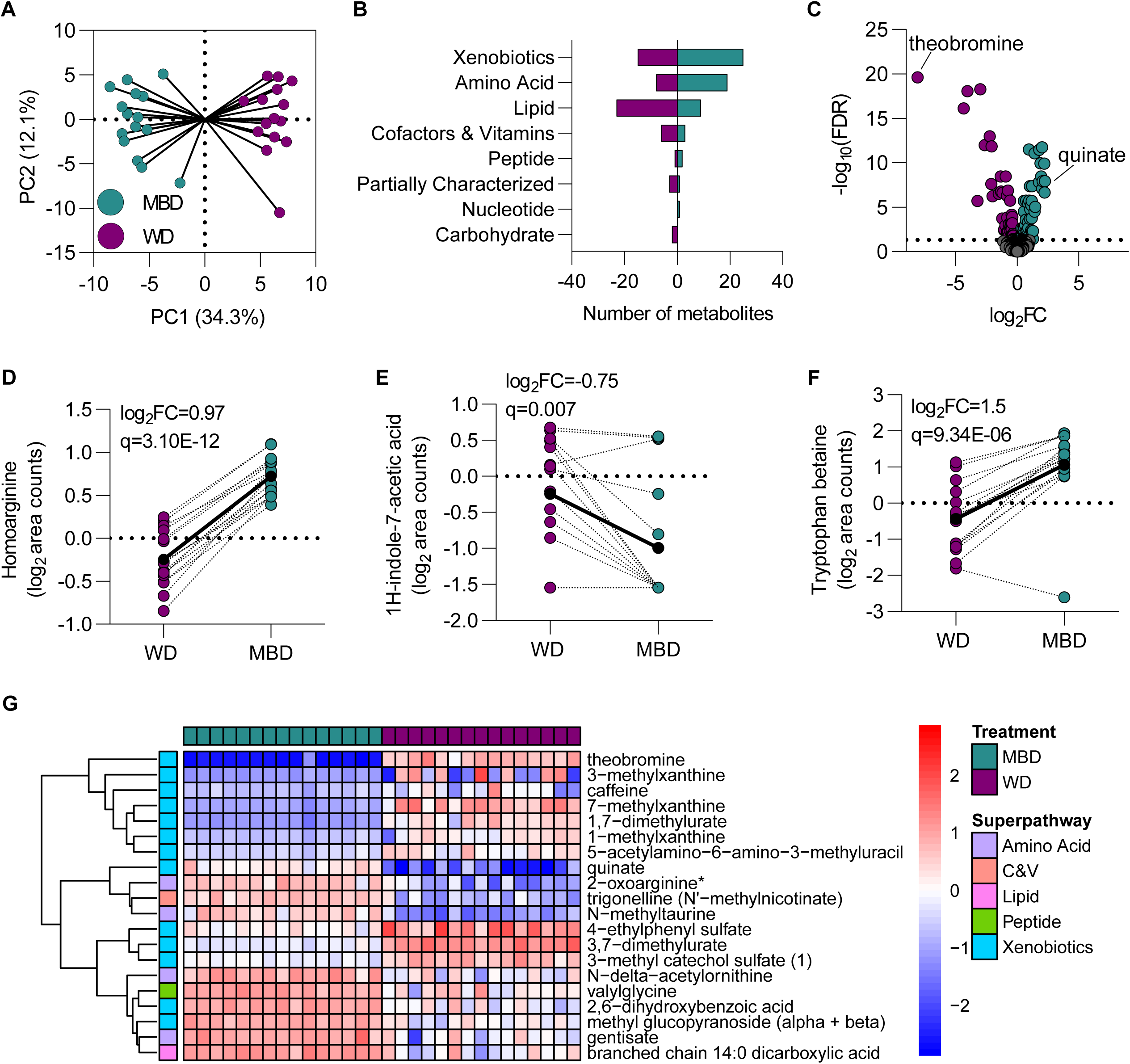
Host-diet-microbiome interactions characterize a circulating metabolome that distinguishes an MBD from a WD. **A.** Principal components analysis of the 118 fasting serum metabolites that significantly varied by diet (q<0.05) according to linear mixed effects regression. **B.** Bar plot of serum metabolomic superpathways increased or decreased by an MBD relative to a WD. **C.** Volcano plot showing diet-induced changes among fasting serum metabolites; five metabolites with an absolute log_2_FC >3 were excluded to improve the ease of interpretation. Log_2_ area counts of fasting serum homoarginine (**D.**), 1H-indole-7-acetic acid (**E.**), and tryptophan betaine (**F.**) by diet; log_2_ foldchanges and p-values are from limma mixed effects regression models. **G.** Heatmap of metabolites differentially abundant by diet (q<0.05) with absolute log_2_FC ≥1. Metabolite names ending in ‘*’ or ‘**’ indicate compounds for which a standard is not available but for which there is confidence in the compound identity(70). N=15 participants for all panels. C&V— Cofactors and vitamins; PC—principal coordinate; MBD—Microbiome Enhancer Diet; WD—Western Diet.

To determine which serum metabolites drove the diet separation when considering differentially abundant metabolites and may therefore be biomarkers of diet-induced microbiome remodeling and energy balance, we evaluated metabolite superpathway overrepresentation (**Figure 4B**) and the specific subpathways and metabolites driving these differences. Among the top MBD-enriched metabolites was quinate (log_2_FC 2.3, q=1.16E^-08^), a plant metabolite with antibacterial properties against opportunistic pathogens that protected against diet-induced obesity in rats (37) (**Figure 4C**). Six amino acid metabolites within the urea cycle subpathway, including several microbiota-linked arginine byproducts, were enriched by an MBD. Of these, homoarginine (log_2_FC 0.8, q=2.28E^-13^; **Figure 4D**) can be synthesized by gut microbes from arginine (38), which may protect against obesity by preventing proteolysis and stimulating lipolysis (39). Consistent with prior studies of plant-forward diets (40, 41) or high-fiber interventions (42), N-delta-acetylornithine (40) (log_2_FC 1.9, q=3.43E^-09^) and the microbial tryptophan metabolite tryptophan betaine (41) (log_2_FC 1.5, q=9.34E^-06^; **Figure 4E**) showed robust mean enrichments by an MBD. Conversely, the microbial urea cycle metabolite 1H-indole-7-acetic acid was decreased by MBD feeding (log_2_FC - 0.75, q=0.007; **Figure 4F**), echoing prior literature that this metabolite was decreased by a fiber-rich relative to a protein-rich diet (43). Overall, the pattern of fasting serum metabolites augmented by MBD feeding (**Figure 4G**) may be a hallmark of a high-fiber diet that fuels the negative energy balance phenotypes of microbial biomass growth and fecal energy loss.

Fasting serum metabolites classified diet with 100% accuracy according to a random Forest machine learning algorithm (**Supplemental Figure 4B**), which highlighted MBD-enriched diet-microbiome co-metabolites like quinate and 4-ethylphenyl sulfate among the top discriminating metabolites. 4-ethylphenyl sulfate and 4-vinylphenol sulfate—both microbiota co-metabolites (44) that were top contributors to diet classification—may be hallmarks of positive energy balance and/or highly processed diets. These results are in line with a seminal study conducted by Hall *et al*, in which both of these metabolites were decreased in the plasma and urine following two weeks of a controlled, minimally processed diet relative to an ultraprocessed diet fed in a domiciled ward. Under these experimental conditions, the ultraprocessed diet increased ad libitum calorie consumption and weight gain (45); thus, 4-ethylphenyl sulfate and 4-vinylphenol sulfate may be biomarkers of a highly processed Western Diet that maximizes energy absorption. In sum, fasting serum metabolite profiles may be useful as biomarkers of the gut microbiota-mediated biotransformation of host and dietary substrates in the context of controlled diets that have disparate effects on energy balance.

### Shared fecal and serum metabolites are marked by host-microbiota co-metabolism

It is well established that microbial metabolites cross the colonic epithelium and enter host circulation via portal vein system both by passive and active transport mechanisms (46). Thus, we next turned our attention to reconciling the feces and fasting serum, as blood samples are collected more routinely in clinical studies and could therefore improve translatability to larger populations. The directionality of metabolite changes was consistent for 22 of 24 metabolites that differed by diet in both feces and fasting serum (**Figure 5**), indicating that levels of fecal metabolites are directly related to circulating levels. Overall, the metabolites altered by MBD compared to WD feeding in both the feces and fasting serum reflected changes in diet-host-microbiota metabolism. This included MBD-induced increases in quinate from tryptophan metabolism, decreased 3-methylhistidine, a gut microbiota metabolite of amino acid degradation (47), and increased trigonelline (N’-methylnicotinate), a plant NAD metabolite that serves as an energy source for gut microbes (48). In addition, we observed beneficial changes in fatty acid metabolites on an MBD, including decreased acyl carnitines, which are increased in the plasma of persons with obesity (49). Several xanthine metabolites were decreased by an MBD, which could reflect either caffeine or theobromine degradation. Dietary theobromine was lower on an MBD due to chocolate-containing food items provided on a WD and not an MBD, although both diets were low in caffeine (<6 mg per ∼2000 kcal per day). Collectively, the 21 metabolites that were not driven by unintended diet differences in caffeine and theobromine may serve as biomarkers of an MBD. Pending confirmation in targeted assays and in larger sample sizes, these metabolites may be useful for verifying dietary adherence and metabolic response to high-fiber, minimally processed diets using blood samples when rigorous fecal sampling is infeasible.

**Fig. 5:**
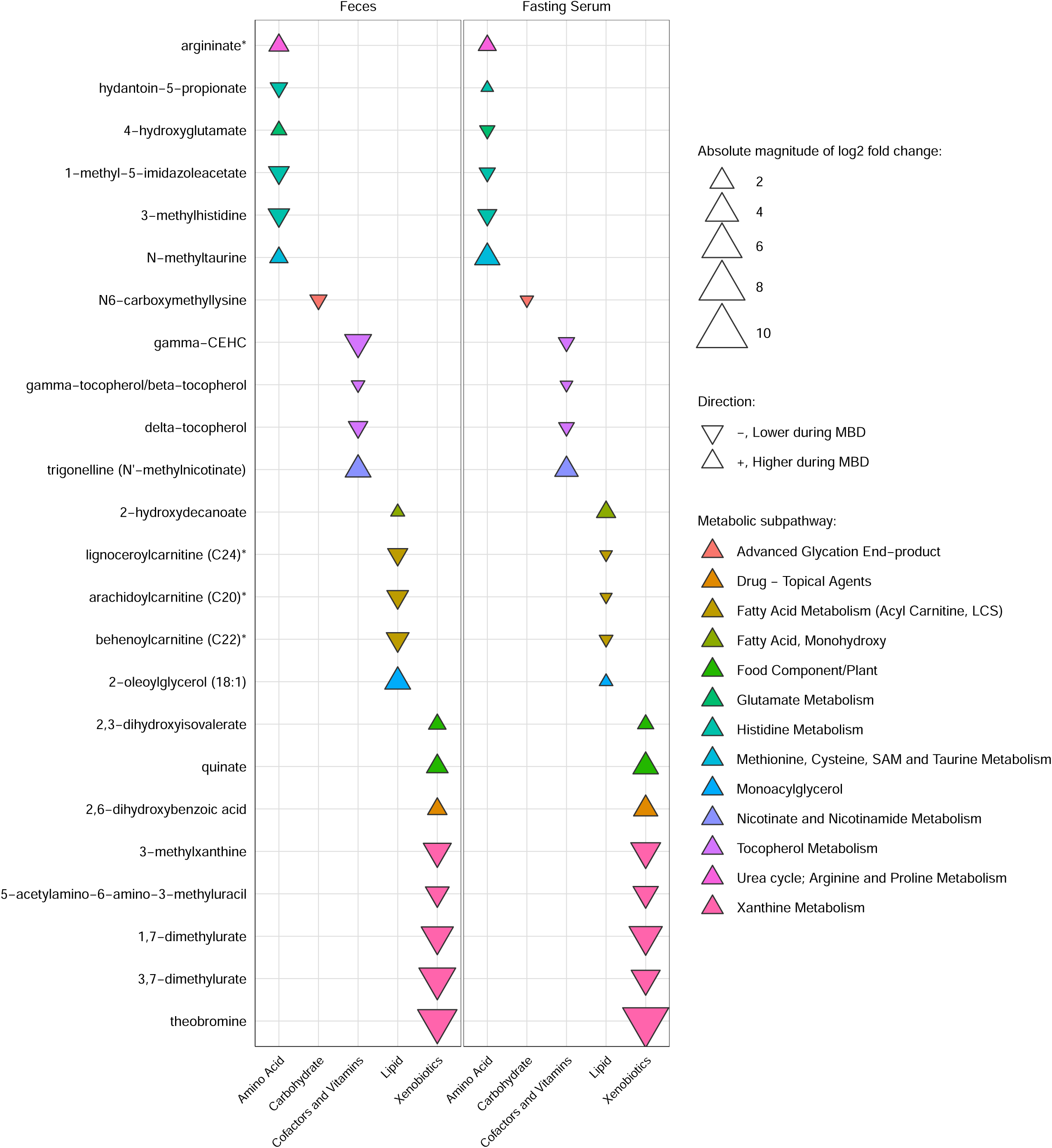
Shared fecal and serum metabolites are marked by host-microbiota co-metabolism. The 24 metabolites altered by MBD relative to WD feeding in both the feces and fasting serum according to limma linear mixed models with participant as random effect and diet, period, and sequence as fixed effects. N=17 participants for fecal data and N=15 participants for fasting serum. Metabolite names ending in ‘*’ or ‘**’ indicate compounds for which a standard is not available but for which there is confidence in their identity(70). PC—principal coordinate; MBD—Microbiome Enhancer Diet.

### An MBD shifts the urinary metabolome from simple sugar degradation to ketogenesis

To garner additional insights into integrated host-microbiota responses to our two experimental diets, we explored the 24-hour urinary metabolome. Similar to the feces, the urinary metabolome showed clear separation by diet whether restricting to the 495 metabolites that varied by diet (q<0.05; **Figure 6A-6B**) or including all 899 metabolites retained after filtering procedures (**Supplemental Figure 5A**). We observed notable decreases in the energy and carbohydrate superpathways on an MBD (**Figure 6C-D**), similar to the feces (**Figure 2B**). Broad decreases in urinary metabolites from the carbohydrate and energy superpathways and the TCA cycle subpathway, alongside increased urinary ketones, served as robust indicators of negative energy balance during MBD relative to WD feeding. A hallmark of these indicators of negative energy balance was a decrease in 21 urinary metabolites of carbohydrate metabolism during MBD relative to WD feeding. This included decreased simple monosaccharides such as fructose (log_2_FC -1.2, q=4.57E^-05^), disaccharides such as lactose (log_2_FC -0.73, q=0.0003), metabolites related to glycolysis and gluconeogenesis such as glycerate (log_2_FC -1.9, q=8.78E^-18^) and lactate (log_2_FC -0.71, q=1.36E^-06^), and pentose sugars such as xylose (log_2_FC -1.4, q=1.15E^-11^) and ribitol (log_2_FC -0.30, q=0.01). Similar to the fecal and fasting serum results, there was a robust MBD-induced decrease in the advanced glycation endproduct N6-carboxymethyllysine (log_2_FC -1.5, q=3.43E^-15^; **Figure 6E**) from carbohydrate metabolism. The exception was an increase in the sugar acid galactonate (log_2_FC 1.0, q= 1.23E^-06^; **Figure 6F**), which is a byproduct of host and microbial metabolism and a carbon energy source for enteric bacteria that facilitates gut microbiota colonization (50). These results indicate a shared metabolomic signature of reduced simple carbohydrate degradation and negative energy balance across the feces, fasting serum, and urine that may impact gut microbial biomass growth and thus host energy absorption.

**Fig. 6:**
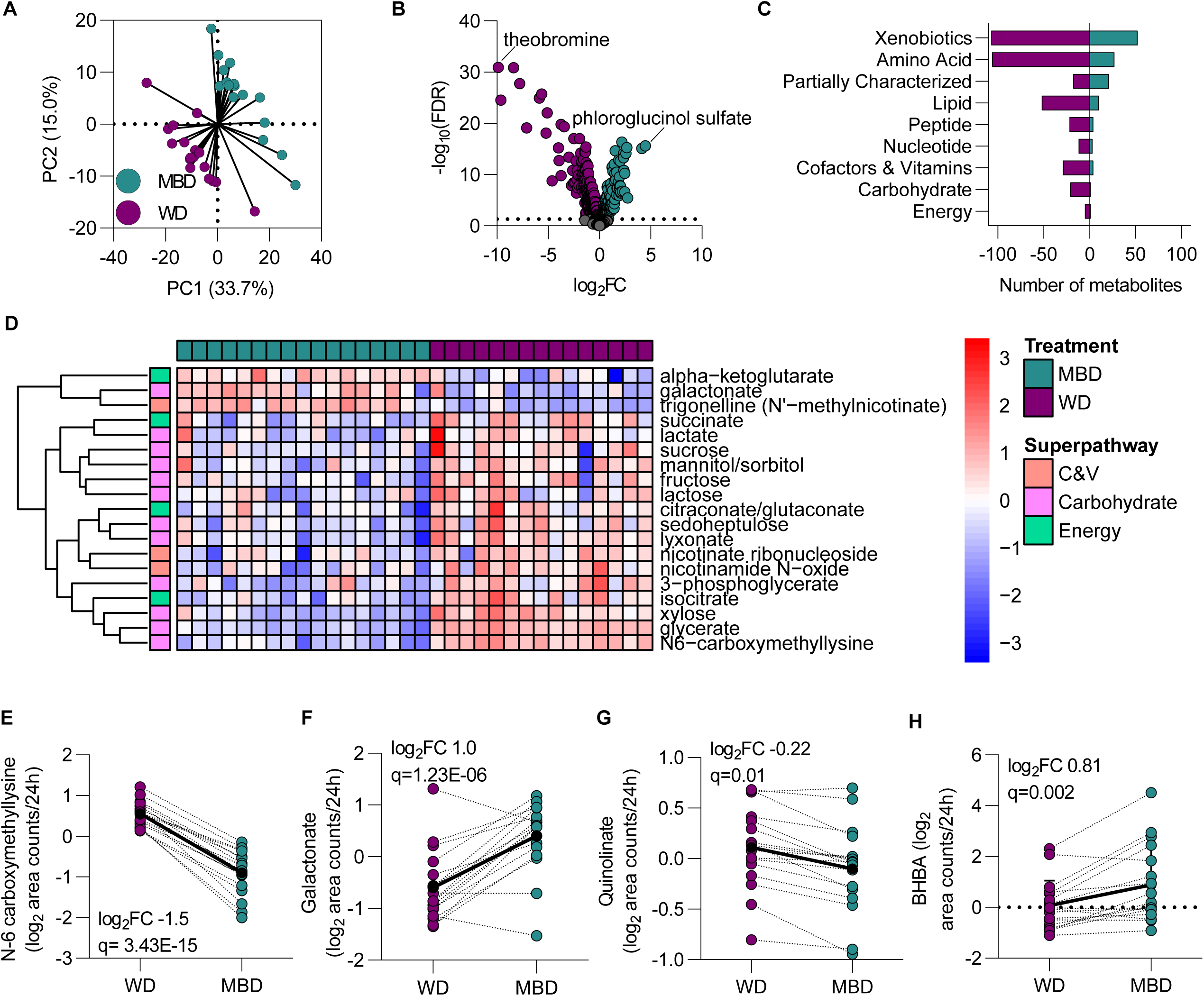
An MBD shifts the urinary metabolome from simple sugar degradation to ketogenesis. **A.** Principal components analysis of the 495 urinary metabolites that significantly varied by diet. **B.** Volcano plot of log_2_ fold-changes in urinary metabolites. **C.** Bar plot of urinary metabolite superpathways enriched and decreased by an MBD. **D.** Heatmap of carbohydrate, energy, and NAD-related metabolites that varied by diet according to limma linear mixed models (q<0.05). **E-F.** Key carbohydrate (**E-F.**), and NAD (**G**) metabolites altered by an MBD relative to a WD (q<0.05). **H.** Urinary 3-hydroxybutyrate (BHBA) was increased by an MBD. Metabolites included in **A-H.** based upon limma linear mixed models with participant as random effect and diet, period, and sequence as fixed effects. N=17 participants for all panels. Metabolite names ending in ‘*’ or ‘**’ indicate compounds for which a standard is not available but for which there is confidence in the compound identity (70). 5-a-6-a-3-mu—5-acetylamino-6-amino-3-methyluracil; C&V—Cofactors and vitamins; PC—principal coordinate; MBD— Microbiome Enhancer Diet; WD—Western Diet; NAD—Nicotinamide.

Complementing the decrease in urinary metabolites of simple carbohydrate degradation was a decrease in five NAD metabolites. Nicotinate ribonucleoside (log_2_FC -0.51, q= 6.78E^-05^), nicotinamide N-oxide (log_2_FC -0.56, q=9.72E^-05^), and quinolinate (log_2_FC -0.22, q= 0.01; **Figure 6G**) were all decreased during MBD relative to WD feeding, with the decrease in quinolinate echoing fecal results. In concert with the decrease in carbohydrate metabolites, an MBD decreased levels of the energy metabolite phosphate (log_2_FC -0.25, q=0.007) and the TCA cycle metabolites isocitrate (log_2_FC -0.77, q=2.94E^-07^), succinylcarnitine (C4-DC; log_2_FC -0.30, q=0.01) succinate (log_2_FC -0.55, q=3.03E^-07^), and citraconate/glutaconate (log_2_FC-0.76, q=4.42E^-05^). The decreased citraconate/glutaconate was similarly reflected in the feces. Again confirming the fecal results, MBD feeding enriched the keystone TCA metabolite alpha-ketoglutarate (log_2_FC 1.3, q=1.28E^-07^), which is critical for gastrointestinal energy metabolism (28). We hypothesize that these urinary metabolite alterations reflect a reduced energy availability to the host when the gut microbiota are adequately fed on an MBD. In other words, an MBD-augmented microbial biomass exploits the available carbon sources in the colon for its own energetic needs, thus preventing absorption of these carbohydrate and energy metabolites across the gut epithelium and shunting energy away from the host.

Accompanying the decrease in simple carbohydrate metabolites, an MBD increased the urinary ketone beta-hydroxybutyrate (BHBA; log_2_FC 0.81, q=0.002; **Figure 6H**). This suggested an increased fat oxidation with downstream ketogenesis on an MBD relative to a WD. These changes occurred despite strict eucaloric and isocaloric dietary conditions. Similar to the feces and serum, a random Forest model predicted diet condition from urinary metabolites with 100% accuracy when including those metabolites that varied by diet (q<0.05; **Supplemental Figure 5B**). Taken together with the metabolites that were robustly altered between diets (**Supplemental Figure 5C**), these metabolites represent a potential biomarker signature of an MBD integrated over 24 hours of urine collection. Prominent in these signatures are connections to gut microbial metabolism, including the MBD-enriched amino acid derivatives N-methylleucine and N-methyltaurine, and the peptide valylglycine, which can be produced by certain soil microbes (51). We conclude that key interactions amongst the gut microbiota and host metabolism influence nutrient processing, leading to reduced host energy absorption and negative energy balance on an MBD.

## Discussion

Our work capitalizes on an exquisitely controlled feeding paradigm with two diets vastly different in microbiota-accessible substrates to identify potential metabolic mechanisms by which diet-driven gut microbiome remodeling drives comprehensive and precise measurements of human energy balance (5). Our work adds to the preclinical literature suggesting a causal role of the gut microbiome in energy balance that is just beginning to emerge in human studies (3, 5, 52).In so doing, we present two major findings. First, we identified robust metabolomic signatures of host energy balance across multiple biological samples that are driven by diet-host-gut microbiome interactions. This includes the decrease in numerous fecal amino acid metabolites during MBD relative to WD feeding likely attributable to the shift in microbial metabolism towards preferred fiber fermentation and away from less energy efficient amino acid fermentation (53). In addition, two microbiota-linked mucin metabolites were decreased by MBD feeding. Mucin degrading gut microbes are known to bloom under conditions of fiber scarcity as the result of feeding on host mucins when dietary fiber is scarce, and they were accordingly decreased by an MBD (5). These signatures serve as putative biomarkers of a “starved” gut microbiome diverting dietary energy to the host (WD) vs. a “fed” gut microbiome (3, 4, 54) that is associated with reduced host energy absorption (MBD). Secondly, we generated hypotheses regarding the molecular mechanisms underlying the negative energy balance induced by an MBD relative to a WD.

Obesity is a disease of positive energy balance; several of the host-microbiota co-metabolites decreased by an MBD have been linked to human obesity in published literature. This includes 3-methylhistidine, which emerged as a hallmark fecal metabolite of positive energy balance. 3-methylhistidine has been associated with higher BMI or obesity in three independent cohorts (58, 59) and decreased ∼180-fold by an MBD in the feces. An MBD-induced decrease in fecal quinolinate, an NAD precursor linked to microbiota metabolism of tryptophan and kynurenine, was correlated with increased energy absorption and decreased microbial biomass in our data. This finding is supported by a previous correlation of quinolinate with higher BMI in women with obesity (60). An MBD also decreased fecal P-cresol, a proposed microbial uremic toxin directly associated with obesity in a meta-analysis of thirty articles (61). The ability of p-cresol to feed a microbial biomass capable of fermenting simple carbohydrates positions this metabolite as a biomarker of positive energy balance and/or low fiber intake that can be decreased by an MBD. Pathway analysis of fecal metabolites revealed that one mechanism by which an MBD may induce negative energy balance is through activation of PPARα—a gene which stimulates lipid catabolism and is associated inversely with obesity in experimental animal (32) and human models (33). Intriguingly, the gut symbiont *Parabacteroides distasonis*—whose relative abundance was increased by an MBD (5)—augmented lipid catabolism in mice via PPARα activation (34). Taken together with the restructuring of fecal lipid composition by an MBD, these results suggest that fiber-induced microbiota remodeling may reduce host energy absorption through mechanisms related to lipid handling.

Whereas metabolites decreased by an MBD have been associated positively with obesity, several of the metabolites increased by an MBD have shown protective correlations countering obesity. These metabolites include 1,2-dilinoleoyl-GPC (18:2/18:2), which showed among the greatest MBD-induced increases in the feces and has been associated inversely with obesity in a large observational cohort study (62). Similarly, urinary enterolactone produced via microbial lignin degradation (30) was enriched by an MBD ∼4-fold and was associated negatively with obesity in a population-based study (31). This signal may be the direct result of reduced obesity-associated gut microbial species such as *Clostridium leptum and Clostridium bolteae*. The relative abundance of these two species were decreased by an MBD (5) and have been correlated negatively with plasma enterolactone in a large observational study (30).

Thus, enterolactone may mediate the effects of diet-induced gut microbiome remodeling on energy balance. Fecal Urolithin A was increased by an MBD ∼64-fold. In experimental literature, greater urinary Urolithin A was related to increased visceral fat mass loss in 294 human participants provided with polyphenol-rich foods (22). In summary, prior literature corroborates that numerous microbiome-linked metabolites altered by an MBD have been correlated with the energy balance phenotype of obesity. Upon replication, future studies are needed to determine which microbes and metabolites are quantitatively important for energy balance.

Urinary metabolite signatures have been used to study the metabolic impacts of gut microbiota remodeling (63). Here, we harnessed 24-hour urinary metabolomics data in the context of an energy balance paradigm. Our global urinary signatures uncovered that feeding an MBD increased urea cycle and TCA metabolites and the ketone body BHBA. These findings raise the possibility that an MBD optimizes the efficiency of energy metabolism through the TCA and urea cycle (64), thereby mimicking fasting conditions to induce ketogenesis without intentional energy restriction or changes in macronutrient content. We surmise that gut microbiota remodeling contributed to this effect. One indicator supporting this hypothesis was a decreased relative abundance of several *Bifidobacterium* species in our published results (5), as reductions in specific species within this genera have been associated with increased plasma ketones in a domiciled randomized crossover study (65). Future studies should investigate the hypothesis that decreasing the relative abundance of certain *Bifidobacterium* species, among other relevant members of the gut microbial community, may release inhibitory brakes on fatty acid oxidation and ketogenesis.

In addition to the advancements in our understanding of the impact of controlled microbiota-focused diets on metabolomics profiles and energy balance, we want to highlight one other intriguing set of observations. Over two-thirds of the fecal and urinary small molecules we detected were decreased by an MBD. Several hypotheses may explain this finding. First, only a small proportion of the total metabolome is detected by existing metabolomics platforms (55). Furthermore, the apparent global decrease in metabolites on an MBD could be an artifact of existing spectral libraries being biased towards biomolecules of human origin, as most microbial molecules are not commercially available to be used as chemical standards (56). Alternatively, it is possible that MBD-enriched diet microbiota co-metabolites were readily taken up by bacterial cells and used to synthesize macromolecules (i.e., DNA, carbohydrates, etc.) required to form the structure of microbial cells and for energy production (57). Given that such macromolecules are not detected by global metabolomics assays, methods that detect these molecular structures and differentiate microbial from host macromolecules are required to test this hypothesis. A complementary hypothesis is that the decreased microbial biomass on a WD leads to incomplete degradation of dietary substrates that reach the colon, leading to more end products of “incomplete” substrate utilization. In theory, a robust MBD-augmented microbial biomass engages in more extensive cross-feeding (57), harnessing all small molecules available for biomass growth. Thus, metabolites that “feed” microbial biomass may protect against obesity by reducing host energy absorption.

Our controlled feeding energy balance paradigm has both strengths and weaknesses that are worthy of acknowledgement, in addition to those already mentioned. The domiciled controlled feeding design allowed for causal inference regarding the effects of our two diets on the human metabolome while circumventing the reporting bias inherent to self-reported dietary intake. The crossover design minimized intra-individual variation and eliminates the impact of numerous potential confounders, allowing for detection of unbiased group differences by diet. Metabolomic signatures measured in the feces, fasting serum, and 24-hour urine allowed for verification of metabolite changes induced by our experimental diets. Our robust analytic pipeline uncovered potential mechanisms linking diet-microbiota co-metabolites to energy balance phenotypes. Despite these strengths, our study design did not allow for direct estimation of the effects of microbiome remodeling on the metabolome, or vice versa. Despite an exquisitely controlled crossover experimental paradigm minimizing within-and between-participant noise, the modest sample size may have precluded detection of metabolite changes or correlations with clinical traits in some cases. As mentioned previously, metabolomics platforms do not detect macromolecules, nor measure metabolic flux. Thus, future studies should evaluate the contribution of small molecules to microbial biomass growth and their flux from the colon into circulation and urinary excretion. Furthermore, the impact of metabolites on microbial composition can have direct effects on microbial biomass (66)—a key energy balance phenotype (5). Thus, bidirectional interactions amongst diet, metabolites, and the gut microbiota should be examined to gain a clearer picture of microbial mechanisms mediating the impact of diet on host energy absorption and energy balance.

Our comprehensive evaluation of metabolomic signatures differentially altered by MBD and WD feeding revealed potential mechanisms by which a high-fiber whole foods diet reduces dietary energy absorption and increases microbial biomass—a salient component of fecal energy loss. Although the focus of our study was energy balance, many of the microbial co-metabolites we identified have been linked either positively or negatively with cardiometabolic disease including type 2 diabetes and insulin resistance (23, 61, 67–69), and may therefore be useful for studying host-diet-microbiome interactions related to the downstream sequalae of positive energy balance in future studies. This initial glimpse into global metabolomic signatures should be replicated in larger studies and with targeted assays to identify therapeutic mechanisms by which host-diet-microbiome interactions can help curb the obesity epidemic.

## Acknowledgements

We thank our study participants, without whom this work would not have been possible. Research reported in this publication was supported by the National Institute of Diabetes and Digestive and Kidney Diseases of the National Institutes of Health under Award Number R01DK105829. The content is solely the responsibility of the authors and does not necessarily represent the official views of the National Institutes of Health.

## Author contributions

DI, KDC, SRS, and GY designed research. GY and DI performed the bioinformatic and statistical analyses. KDC, SRS, RK-B and EC participated in the design of the parent clinical trial. DI wrote the initial draft of the manuscript. All authors critically reviewed the manuscript, provided feedback, and agree with the content. SRS and RK-B secured grant funding.

## Data availability

Data, code book, and analytic code will be made available upon request pending approval from the study investigators. The R code for the limma package used for linear mixed models, the WGCNA package used for network analysis with fecal metabolites, and the randomForest package used to predict diet assignment from metabolites is publicly available in a GitHub repository: https://github.com/dadaria89/MAC/blob/09308aef7cc287811673c3b75d77ebc7dfc92ad1/Metabolomics.

## Abbreviations

BMI: body mass index
CTT: colonic transit time
MBD: Microbiome Enhancer Diet
NAD: nicotinamide
PP: Pancreatic polypeptide
PCA: principal components analysis
UPLC-MS/MS: Ultrahigh Performance Liquid Chromatography-Tandem Mass Spectroscopy
WD: Western Diet
WGCNA: weighted gene correlation analysis

---

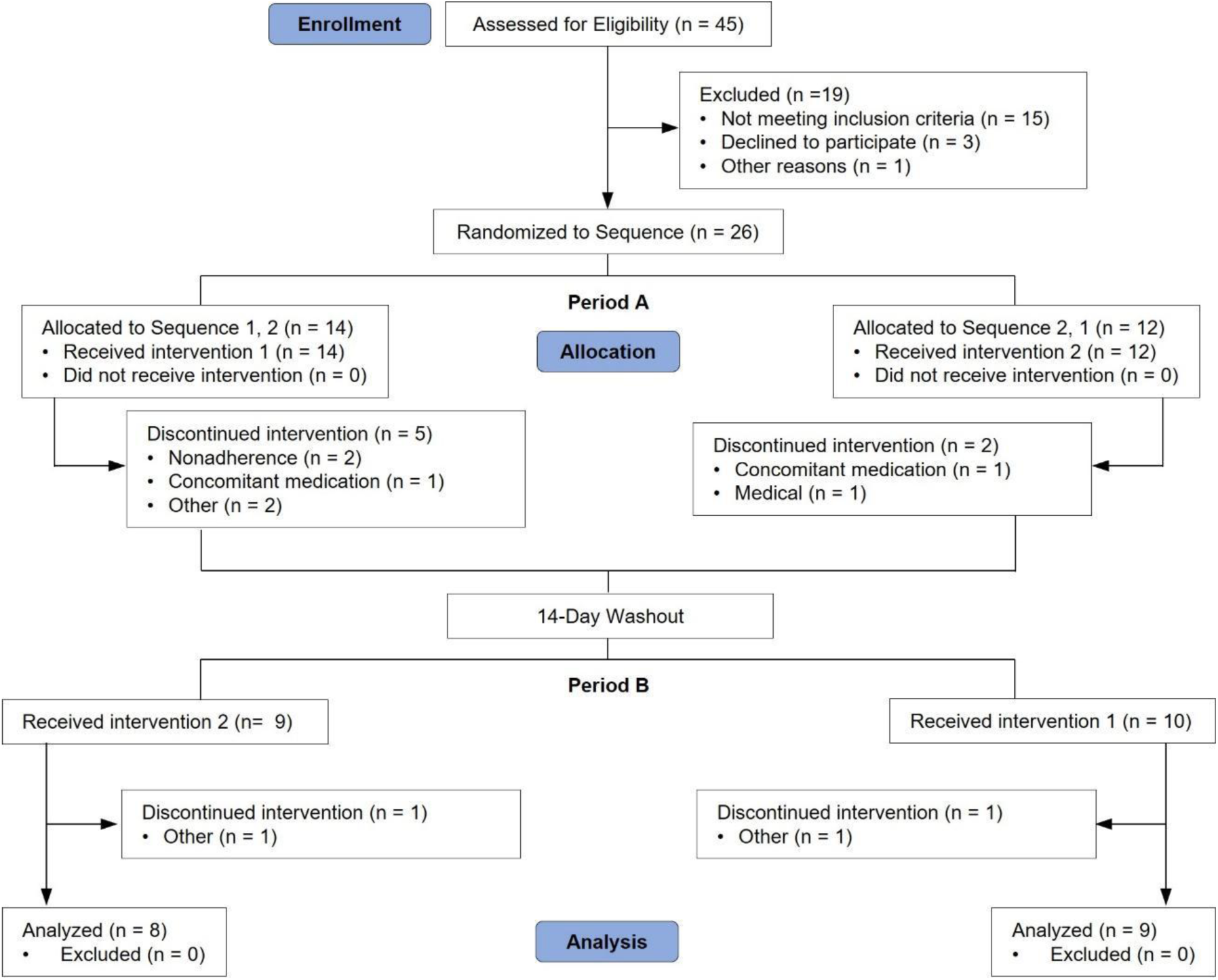

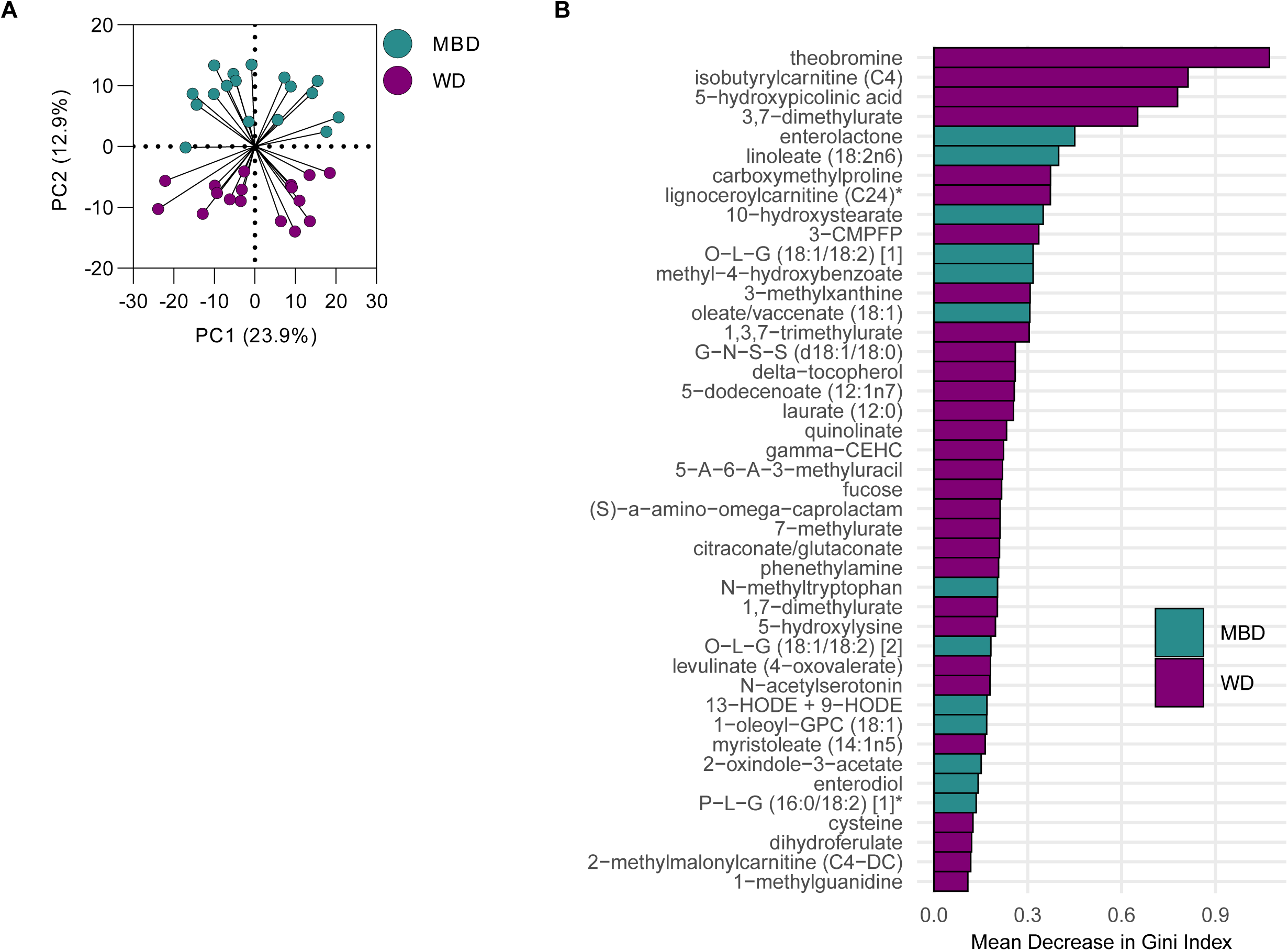

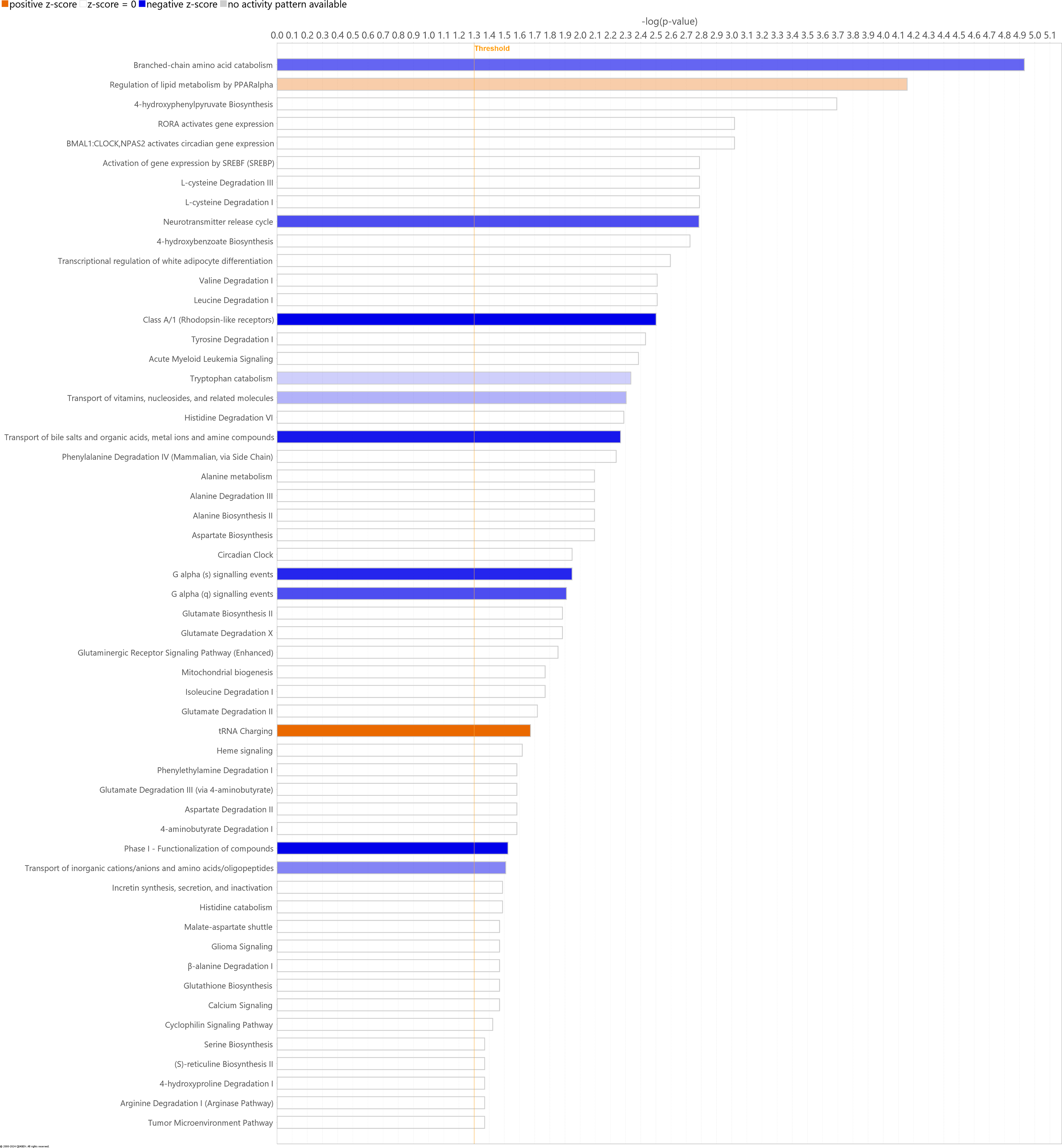

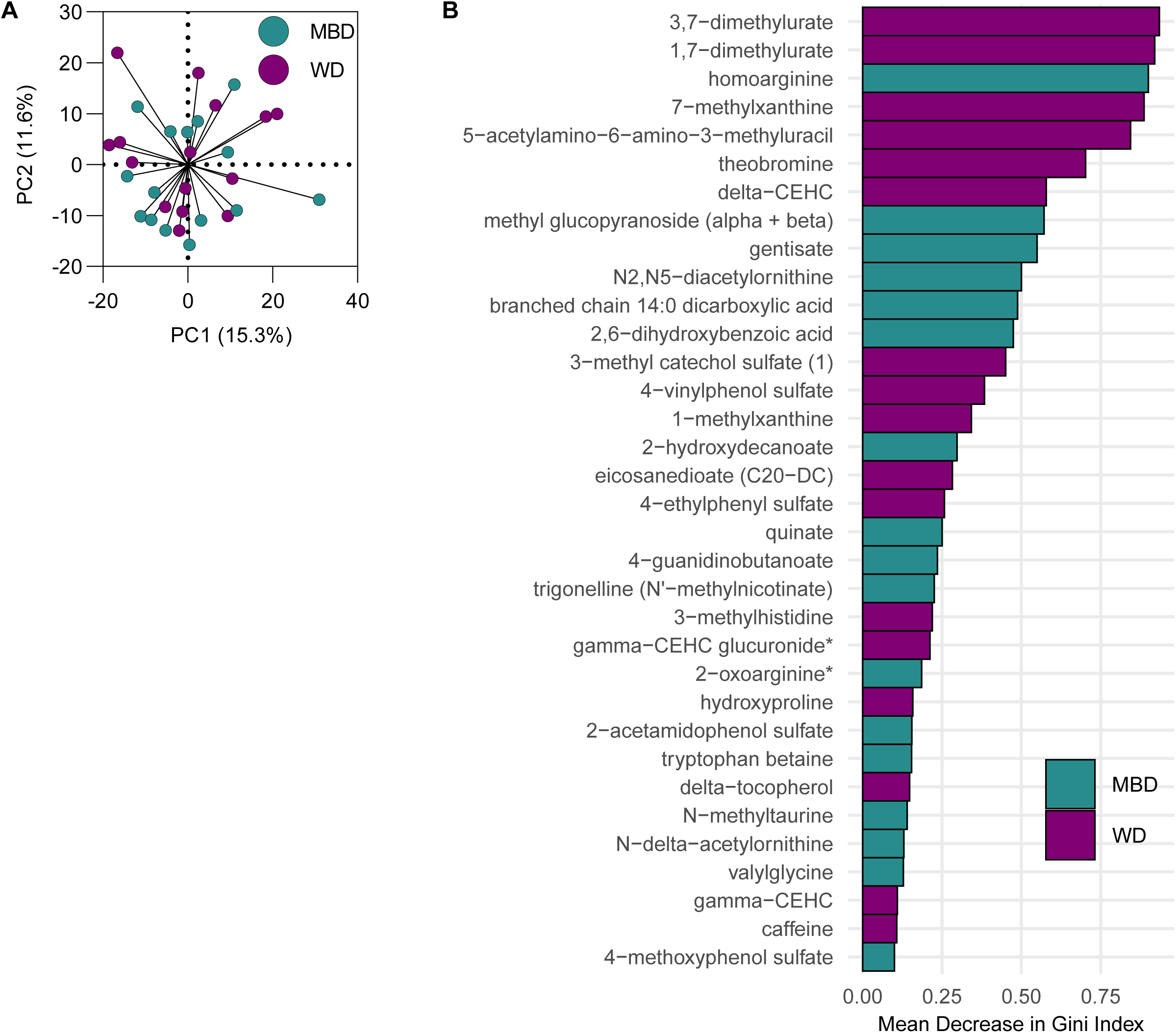

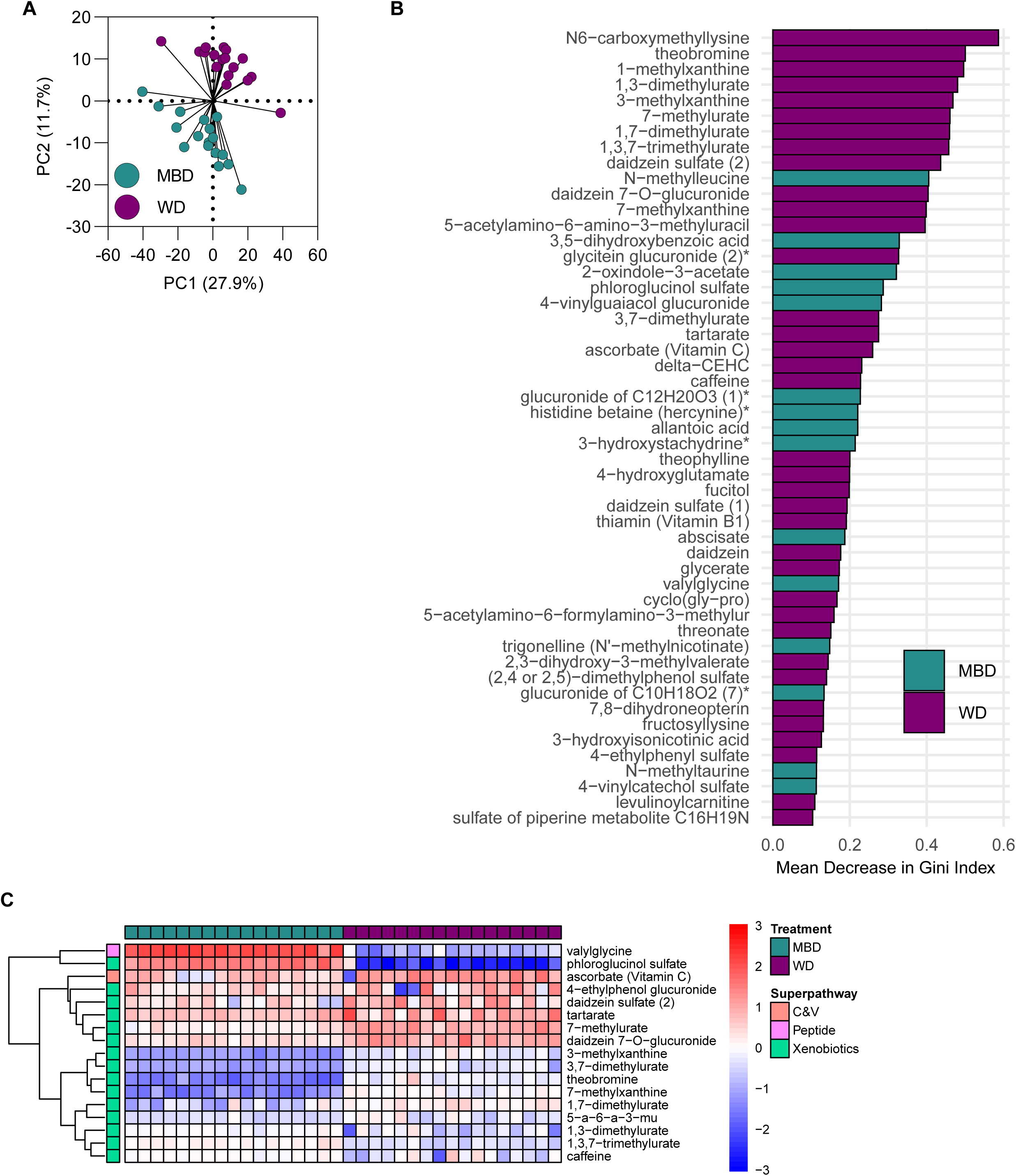

